# Post Tuberculosis (TB) Bronchiectasis versus Non-TB Bronchiectasis in Northern Pakistan: A single centre retrospective cohort study on frequency, demographics, microbiology, and complications

**DOI:** 10.1101/2024.02.19.24303057

**Authors:** Anbesan Samuel Hoole, Ahsan Ilyas, Sunaina Munawar, Matthew Cant, Rizwan Hameed, Shahzad Gill, Joel Riaz, Issac Siddiq

## Abstract

**Introduction:** While Post Tuberculous (TB) Bronchiectasis is the most common cause of Bronchiectasis in South Asia, there has been little research into its microbiology and clinical characteristics in Pakistan. This single centre retrospective cohort study at Bach Christian Hospital (BCH) in rural Northern Pakistan seeks to address this issue.

**Methods:** Demographic, Imaging, Aetiological and Microbiological data were obtained from 32 patients with Bronchiectasis at BCH from between January 2023 and December 2023(1–3).

**Results:** 76% (25/32) of all cases of Bronchiectasis were Post TB. TB infection was seen in 5 cases of Post TB Bronchiectasis, TB with bacterial or fungal co infections in 4, and single bacterial infections in 4. In post TB Bronchiectasis 4 patients had growth of a single bacterium. Drug sensitivities were obtained for bacterial isolates.

One patient each with Post TB and Non-TB Bronchiectasis died from Type 2 Respiratory failure despite appropriate treatment. 2 patients with Post TB Bronchiectasis and destroyed lung syndrome improved but with ongoing significant respiratory impairment(4). All other patients improved with treatment.

**Discussion:**

1. The frequency of Post TB Bronchiectasis is very high even for South Asia(5–8).
2. A significant number (8/24) of Post TB Bronchiectasis had re-infection or failure to improve despite appropriate drug treatment. TB PCR (Polymerase Chain Reaction) on Bronchoalveolar lavage (BAL) was key in the management of these patients(9).
3. Among patients with Post TB Bronchiectasis, those with co-infection present a difficult treatment challenge.
4. Some patients with Post TB Bronchiectasis have significant complications such as destroyed lung syndrome which is difficult to manage.
5. Drug susceptible bacteria and NTM were less commonly isolated than in other studies(10–12).

**Conclusion:** Further research is needed particularly to manage Post TB Bronchiectasis patients with co-infections or complications such as significant structural lung disease.

**Key Messages:** *What is known about this topic:* Tuberculosis (TB) is the most common cause of bronchiectasis in South Asia, but little is known about microbiology or clinical characteristics of post-TB Bronchiectasis.

*What this study adds:* Post TB Bronchiectasis has a very high prevalence in our region, with TB PCR key for diagnosis of TB re-infection in some cases. Mycobacterial, Bacterial and Fungal co-infections form a treatment challenge, as does significant structural lung disease such as destroyed lung.

*How this study might affect research, practice, or policy:* TB PCR testing is a valuable tool in Post TB Bronchiectasis and further research and larger-scale studies are needed to determine optimal treatment for co-infections and significant structural lung disease.

## Introduction

Pakistan has the 5^th^ highest burden of Tuberculosis (TB) world-wide and accounts for 61% of new TB cases in the WHO Eastern Mediterranean Region(13). While a national TB control programme has long existed, there are delays in diagnosis and treatment resulting in a relatively static high incidence of TB(14–17) and increased complications of TB such as bronchiectasis(18). TB is known to be the most common cause of Bronchiectasis in South Asia (7,19) and patients with Post TB Bronchiectasis are known to have more severe disease compared with Non-TB Bronchiectasis (5,20–23). However, there has been little research into its prevalence, microbiology, or complications in Pakistan(8,18). In this single-centre retrospective cohort study at Bach Christian Hospital (BCH) in rural Northern Pakistan, we seek to explore the frequency of TB as a cause of Bronchiectasis, demographics, as well as the microbiology and associated complications in TB versus Non-TB Bronchiectasis.

## Methods

Demographic, Imaging, Aetiological and Microbiological data were obtained from patients with Bronchiectasis at BCH between January 2023 and December 2023. Patients established on TB treatment based on sputum results and improving after two months were not included(1,24). Bronchiectasis was defined according to BTS guidelines based on CT Thorax imaging(3). Aetiology was determined by clinical history, imaging findings and further investigations where feasible. Microbiology was based on Bronchoalveolar lavage (BAL) for all but one patient.

Ethical approval was obtained from BCH ethics committee.

## Results

32 patients with Bronchiectasis were included, out of which 25 (78%) had post TB Bronchiectasis (*figure 1*). Non-TB Bronchiectasis aetiologies included Post childhood infection (n = 2), COPD (n =1), Kartagener’s (n = 1), Rheumatoid Arthritis (n = 1) and Idiopathic (n = 2). The median age in Post TB Bronchiectasis was 65 years (Range 17 – 81), while the median age in Non-TB Bronchiectasis was 45 (Range 14 – 73). There were 15 females and 10 males in the Post TB Bronchiectasis group, and 4 females and 3 males in the Non-TB Bronchiectasis group.

**Figure 1.**
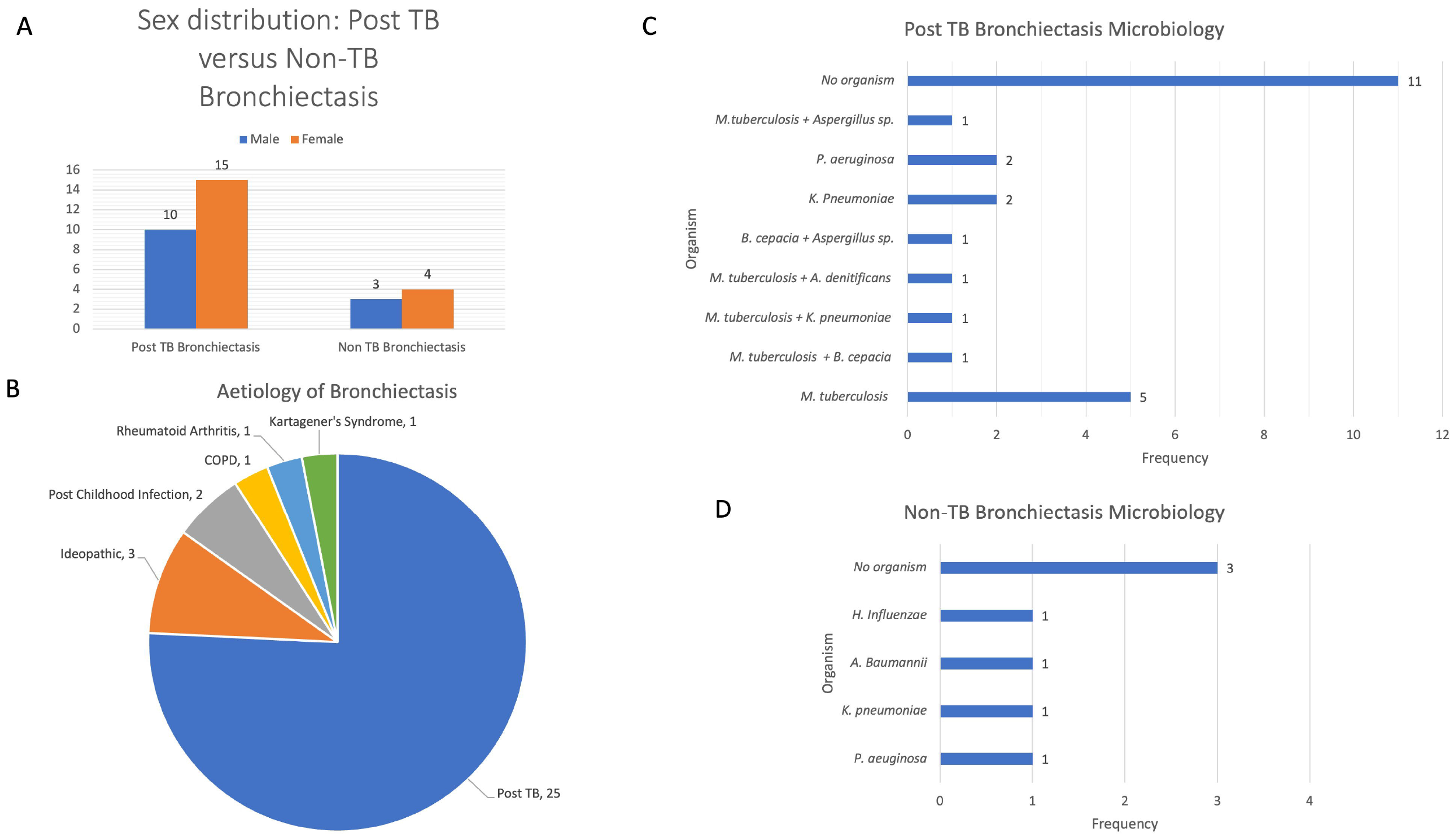
A. Sex distribution in Post TB vs Non-TB Bronchiectasis B. Aetiology distribution of Bronchiectasis C. Microbiology of Post TB Bronchiectasis D Microbiology of Non TB Bronchiectasis

In Post TB Bronchiectasis, 9 patients had *Mycobacterium tuberculosis (M. tb)* on BAL based on Gene Xpert MTB/Rif PCR despite negative sputum Acid Alcohol Fast Bacilli (AAFB). Only one of these patients had AAFB on BAL. No TB patient had Rifampicin Resistance (surrogate for Multi Drug Resistant [MDR] TB). Full drug sensitivities were obtained in two patients which revealed fully drug sensitive (DS) TB.

Four patients with positive TB PCR on BAL had bacterial or fungal co-infections. In addition to *M tb*, one patient each had *Burkholderia Cepacia (B. Cepacia), Achromobacter denitrificans, Klebsiella pneumoniae*, and Aspergillus (BAL Galactomannan positive and imaging suggestive of Pulmonary aspergillosis).

One patient had co-infection with B. cepacia and Aspergillus. Two patients each had Klebsiella and Pseudomonas. In 11 patients with Post TB Bronchiectasis no organism was identified.

In Non-TB Bronchiectasis, one patient each had *Klebsiella pneumoniae, Pseudomonas aeruginosa*, Carbapenemase Resistant *Acinetobacter Baumannii* (CRAB) and *Haemophilus Influenzae (H. Influenzae)*. Three patients had no identifiable organism.

Drug sensitivities were obtained for bacterial isolates (*figure 2*)

**Figure 2.**
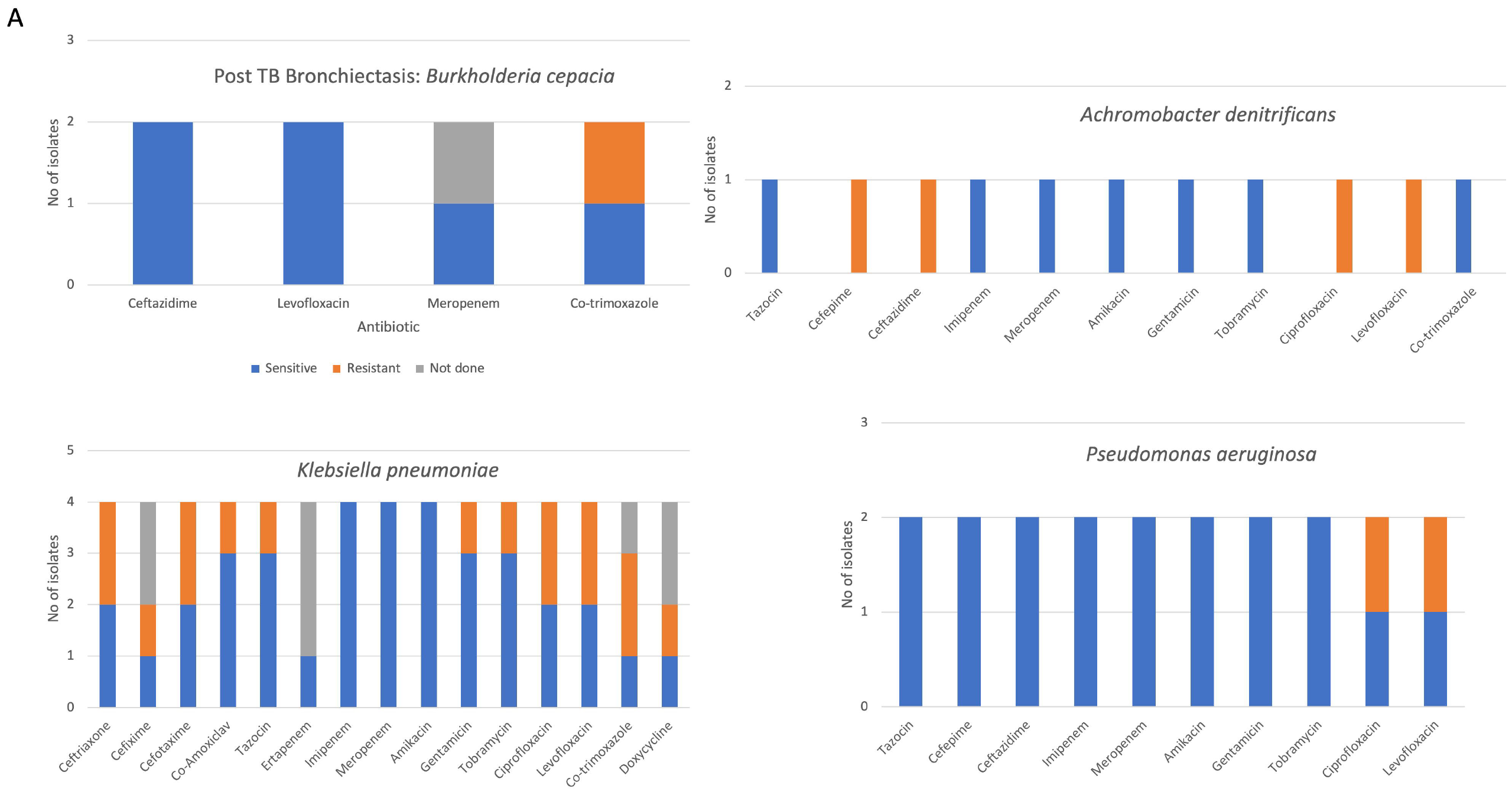
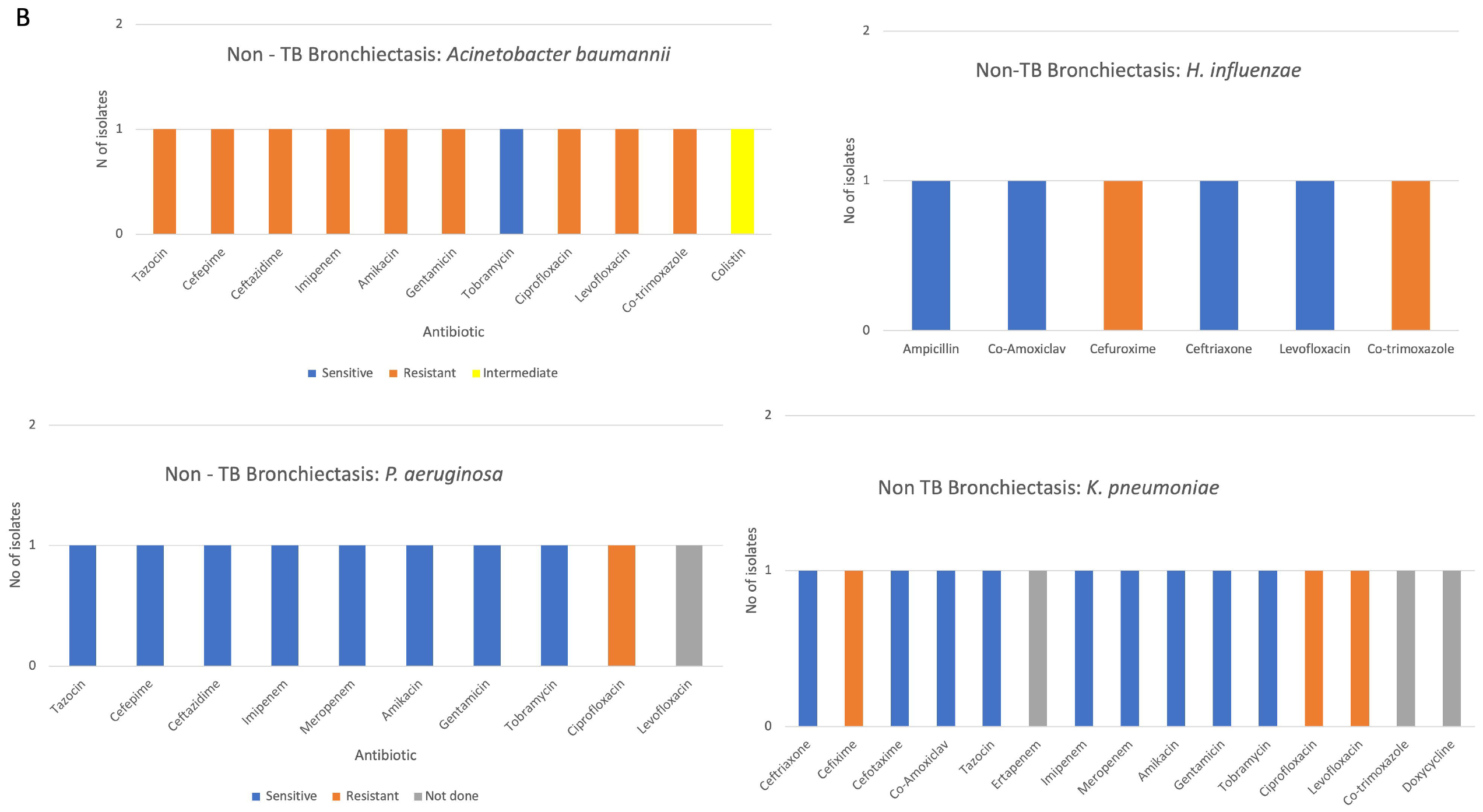
Drug sensitivities of bacterial isolates from Post TB Bronchiectasis (A) and Non-TB Bronchiectasis (B)

In terms of complications, the patient with TB and *B. cepacia* co-infection as well the patient with CRAB died during inpatient admission despite sensitivity directed antibiotic treatment alongside bronchodilators and chest physiotherapy for mucous clearance, as well as eventually NIV for type 2 respiratory failure (T2RF). Two patients with Post TB Bronchiectasis, one with TB infection and the other with *B. cepacia* and Pulmonary Aspergillosis, both presented with destroyed lung syndrome(4) affecting the left lung causing significant functional impairment but have improved with treatment. Even so, both have significant residual respiratory impairment. All other patients have improved with treatment.

## Discussion

Firstly, the frequency of Post TB Bronchiectasis (76% of all Bronchiectasis cases) is very high compared with other studies, even for South Asia. Notably a previous study from Southern Pakistan reported 58% and an Indian study reported 50%(7,8). This is higher than other areas of the world, even in Asia (Saudi Arabia 43%, Korea 20%)(5,6).

Secondly, a significant number (8/24) of Post TB Bronchiectasis patients had re-infection with TB or failure to clinically improve despite an adequate period of appropriate TB treatment (Range: 2 months – 1 year). Notably none of these patients were sputum AAFB positive, highlighting the diagnostic challenge of paucibacillary TB. This is an important clinical problem in cases of TB reinfection, where the diagnosis may be missed in rural TB centres utilising only sputum AAFB. This can be largely overcome by sputum MTB/Rif PCR available in some of the secondary care centres(9), however one 14-year-old male diagnosed with TB reinfection from BAL PCR had previous negative sputum and gastric lavage PCRs.

Thirdly among patients with Post TB Bronchiectasis, those with co-infection present a difficult treatment challenge. The clinical significance of the bacterial and co-infections is uncertain and interactions between M tb and other bacteria and fungi within the microbial interactome are poorly understood(25). In addition, appropriate treatment regimens can be challenging, particularly for drug resistant bacteria such as B. cepacia. Treatment for pulmonary aspergillosis co-infection with TB is also challenging in our setting, due to the drug interactions between rifampicin for TB and antifungal azole drugs. This is compounded by limited access to anti-TB drugs other than the combined fixed dose tablets of either Isoniazid, Rifampicin, Pyrazinamide and Ethambutol (HRZE) or Isoniazid and Rifampicin (HR)(24).

Fourthly, three out of the 25 Post TB patients had significant complications including destroyed lung (n = 2) and death due to T2RF despite NIV. One patient with non-TB Bronchiectasis with CRAB also died due to T2RF despite NIV. Although other studies have highlighted greater disease severity in Post TB Bronchiectasis compared with Non-TB Bronchiectasis, it is difficult to reach any conclusions in this cohort due to small sample size. Significant structural lung disease such as destroyed lung syndrome with large lung cavities is challenging, particularly within our resource-poor context where there is limited recourse to cardiothoracic surgery such as Video-Assisted Thoracic Surgery (VATS) due to cost and availability. Here Endobronchial valve insertion may be an option and has already been trialled in patients with tuberculous cavities(26). While early detection and diagnosis remains key to the reduction of Post TB Bronchiectasis and its complications, further research and resources are also necessary to reduce morbidity and mortality in those who have already developed advanced disease.

Fifthly, organisms such as *H. influenzae* more commonly isolated in other studies were less frequent(10,11). But this likely reflects prior antibiotic treatment eradicating more drug susceptible organisms. Interestingly Non-TB Mycobacteria (NTM) were not isolated in this study either, but this may reflect challenges in diagnostics in Pakistan(12).

Significant limitations of this study include its small sample size and single centre. Statistical tests to determine the significance of differences in terms of microbiology and complications between Post TB and Non-TB Bronchiectasis were not performed due to small sample size. This study could be improved by addition of spirometry and functional status data to complete validated severity scoring systems such as the Bronchiectasis severity index (BSI) (27). A multicentre study for Bronchiectasis in Pakistan could be implemented, like the European Multicentre Bronchiectasis Audit and Research Collaboration (EMBARC) and Respiratory Research Network of India (EMBARC – India) which was recently published(28). This would not only improve understanding of the disease and outcomes for patients in Pakistan, but in a global era also help keep track of pathogens that could spread world-wide.

## Conclusion

Post TB Bronchiectasis is the leading cause of Bronchiectasis in Northern Pakistan, and important yet understudied cause of morbidity and mortality. While Bronchoscopy and advances in technology such as MTB/Rif PCR provide valuable tools in the management of these patients, further research and resources are required particularly in patients with complications such as bacterial or fungal coinfections or significant structural lung disease.

No funding was obtained for this study. The authors declare no competing interests. Patients or the public WERE NOT involved in the design, or conduct, or reporting, or dissemination plans of our research. This study was presented as a poster at the Respiratory Tract Infection Forum in Glasgow in February 2024(29).

## Data Availability

All data produced in the present study are available upon reasonable request to the authors.

## References

1. Treatment of drug-susceptible pulmonary tuberculosis in nonpregnant adults without HIV infection - UpToDate [Internet]. [cited 2024 Jan 11]. Available from: https://www.uptodate.com/contents/treatment-of-drug-susceptible-pulmonary-tuberculosis-in-nonpregnant-adults-without-hiv-infection?search=tuberculosis%20treatment&source=search_result&selectedTitle=1~150&usage_type=default&display_rank=1#H1

2. World Health Organization. WHO consolidated guidelines on tuberculosis. Module 4, Treatment]: drug-resistant tuberculosis treatment.:98.

3. Bronchiectasis in Adults | British Thoracic Society | Better lung health for all [Internet]. [cited 2024 Jan 11]. Available from: https://www.brit-thoracic.org.uk/quality-improvement/guidelines/bronchiectasis-in-adults/

4. Yadav S. Destroyed Lung Syndrome in a Young Indian Male: A Case Report. Cureus [Internet]. 2023 Apr 27 [cited 2024 Jan 11];15(4). Available from: https://pubmed.ncbi.nlm.nih.gov/37252524/

5. Choi H, Lee H, Ra SW, Kim HK, Lee JS, Um SJ, et al. Clinical Characteristics of Patients with Post-Tuberculosis Bronchiectasis: Findings from the KMBARC Registry. J Clin Med [Internet]. 2021 Oct 1 [cited 2024 Jan 11];10(19). Available from: https://pubmed.ncbi.nlm.nih.gov/34640560/

6. Al-Harbi A, Al-Ghamdi M, Khan M, Al-Rajhi S, Al-Jahdali H. Performance of Multidimensional Severity Scoring Systems in Patients with Post-Tuberculosis Bronchiectasis. Int J Chron Obstruct Pulmon Dis [Internet]. 2020 [cited 2024 Jan 11];15:2157–65. Available from: https://pubmed.ncbi.nlm.nih.gov/32982208/

7. Bajpai J, Kant S, Verma A, Bajaj DK. Clinical, Radiological, and Lung Function Characteristics of Post-tuberculosis Bronchiectasis: An Experience From a Tertiary Care Center in India. Cureus [Internet]. 2023 Feb 7 [cited 2024 Jan 11];15(2). Available from: /pmc/articles/PMC9998134/

8. Sharif N, Baig MS, Sharif S, Irfan M. Etiology, Clinical, Radiological, and Microbiological Profile of Patients with Non-cystic Fibrosis Bronchiectasis at a Tertiary Care Hospital of Pakistan. Cureus [Internet]. 2020 Mar 8 [cited 2024 Jan 11];12(3). Available from: /pmc/articles/PMC7138467/

9. WHO | Xpert MTB/RIF: WHO Policy update and Implementation manual. WHO [Internet]. 2016 [cited 2021 Mar 10]; Available from: http://www.who.int/tb/laboratory/xpert_launchupdate/en/

10. Bopaka RG, El Khattabi W, Janah H, Jabri H, Afif H. Bronchiectasis: a bacteriological profile. Pan Afr Med J [Internet]. 2015 [cited 2024 Jan 11];22:378. Available from: /pmc/articles/PMC4796772/

11. Fong I, Low TB, Yii A. Characterisation of the post-tuberculous phenotype of bronchiectasis: A real-world observational study. Chron Respir Dis [Internet]. 2022 Apr 1 [cited 2024 Jan 11];19:1–7. Available from: /pmc/articles/PMC9052827/

12. Karamat A, Ambreen A, Ishtiaq A, Tahseen S, Rahman MA, Mustafa T. Isolation of non-tuberculous mycobacteria among tuberculosis patients, a study from a tertiary care hospital in Lahore, Pakistan. BMC Infect Dis [Internet]. 2021 Dec 1 [cited 2024 Jan 11];21(1). Available from: https://pubmed.ncbi.nlm.nih.gov/33894767/

13. WHO EMRO | Tuberculosis | Programmes | Pakistan [Internet]. [cited 2022 Oct 17]. Available from: https://www.emro.who.int/pak/programmes/stop-tuberculosis.html

14. Fatima R, Haq MU, Yaqoob A, Mahmood N, Ahmad KL, Osberg M, et al. Delivering Patient-Centered Care in a Fragile State: Using Patient-Pathway Analysis to Understand Tuberculosis-Related Care Seeking in Pakistan. Journal of Infectious Diseases. 2017;216:S733–9.

15. Ali SM, Naureen F, Noor A, Fatima I, Viney K, Ishaq M, et al. Loss-to-follow-up and delay to treatment initiation in Pakistan’s national tuberculosis control programme. BMC Public Health [Internet]. 2018;18(1). Available from: https://pubmed-ncbi-nlm-nih-gov.manchester.idm.oclc.org/29523100/

16. Saqib SE, Ahmad MM, Amezcua-Prieto C, Virginia MR. Treatment delay among pulmonary tuberculosis patients within the Pakistan national tuberculosis control program. American Journal of Tropical Medicine and Hygiene [Internet]. 2018 [cited 2021 Mar 10];99(1):143–9. Available from: /pmc/articles/PMC6085810/

17. Incidence of tuberculosis (per 100,000 people) - Pakistan | Data [Internet]. [cited 2021 May 24]. Available from: https://data.worldbank.org/indicator/SH.TBS.INCD?locations=PK

18. Zubair SM, Ali MG, Irfan M. Post tuberculosis radiological sequelae in patients treated for pulmonary and pleural tuberculosis at a tertiary center in Pakistan. Monaldi Arch Chest Dis [Internet]. 2021 [cited 2024 Jan 11];92(1). Available from: https://pubmed.ncbi.nlm.nih.gov/34340298/

19. Chandrasekaran R, Mac Aogáin M, Chalmers JD, Elborn SJ, Chotirmall SH. Geographic variation in the aetiology, epidemiology and microbiology of bronchiectasis. BMC Pulm Med [Internet]. 2018 May 22 [cited 2024 Jan 11];18(1). Available from: https://pubmed.ncbi.nlm.nih.gov/29788932/

20. Dias VL, Canan MGM, Leitão CA, Okuno EA, de Sant’Ana Grd, Miranda JV. Profile of patients with post-tuberculosis bronchiectasis in a tertiary care hospital in Brazil. J Clin Tuberc Other Mycobact Dis [Internet]. 2022 Dec 1 [cited 2024 Jan 11];29:100339. Available from: /pmc/articles/PMC9672944/

21. Kim T, Lee H, Sim YS, Yang B, Park HY, Ra SW, et al. Respiratory symptoms and health-related quality of life in post-tuberculosis subjects with physician-diagnosed bronchiectasis: a cross-sectional study. J Thorac Dis [Internet]. 2021 Aug 1 [cited 2024 Jan 11];13(8):4894–902. Available from: https://pubmed.ncbi.nlm.nih.gov/34527328/

22. Allwood BW, Byrne A, Meghji J, Rachow A, Van Der Zalm MM, Schoch OD. Post-Tuberculosis Lung Disease: Clinical Review of an Under-Recognised Global Challenge. Respiration [Internet]. 2021 Aug 5 [cited 2024 Jan 11];100(8):751–63. Available from: 10.1159/000512531

23. Jordan TS, Spencer EM, Davies P. Tuberculosis, bronchiectasis and chronic airflow obstruction. Respirology [Internet]. 2010 May 1 [cited 2024 Jan 11];15(4):623–8. Available from: https://onlinelibrary.wiley.com/doi/full/10.1111/j.1440-1843.2010.01749.x

24. World Health Organization. WHO consolidated guidelines on tuberculosis: Module 4: Treatment - Drug-susceptible tuberculosis treatment. Geneva: World Health Organization; 2022. PMID: 35727905. 2022;

25. Naidoo CC, Nyawo GR, Wu BG, Walzl G, Warren RM, Segal LN, et al. The microbiome and tuberculosis: state of the art, potential applications, and defining the clinical research agenda. Lancet Respir Med [Internet]. 2019 Oct 1 [cited 2024 Feb 14];7(10):892–906. Available from: http://www.thelancet.com/article/S2213260018305010/fulltext

26. An H, Liu X, Wang T, Liu L, Yan M, Xu J, et al. Endobronchial Valve Treatment of Tuberculous Cavities in Patients with Multidrug-Resistant Pulmonary Tuberculosis: A Randomized Clinical Study. Pathogens [Internet]. 2022 Aug 1 [cited 2024 Feb 14];11(8). Available from: /pmc/articles/PMC9414730/

27. Costa JC, Machado JN, Ferreira C, Gama J, Rodrigues C. The Bronchiectasis Severity Index and FACED score for assessment of the severity of bronchiectasis. Pulmonology [Internet]. 2018 May 1 [cited 2024 Jan 11];24(3):149–54. Available from: https://pubmed.ncbi.nlm.nih.gov/29306672/

28. Dhar R, Singh S, Talwar D, Murali Mohan B V., Tripathi SK, Swarnakar R, et al. Clinical outcomes of bronchiectasis in India: data from the EMBARC/Respiratory Research Network of India registry. Eur Respir J [Internet]. 2023 Jan 1 [cited 2024 Feb 15];61(1). Available from: https://pubmed.ncbi.nlm.nih.gov/36229049/

29. Abstract Book – 6th Forum on Respiratory Tract Infections [Internet]. [cited 2024 Feb 17]. Available from: https://rti-forum.org/?page_id=2051

